# Gamified closed-loop non-pharmacological intervention enhances tic suppression in children

**DOI:** 10.1101/2024.04.17.24303913

**Authors:** Michael S. Rotstein, Sharon Zimmerman-Brenner, Shiri Davidovitch, Yael Ben-Haim, Yuval Koryto, Romi Sion, Einat Rubinstein, Meshi Djerassi, Nitzan Lubiniaker, Tammy Pilowsky Peleg, Tamar Steinberg, Yael Leitner, Gal Raz

## Abstract

**Background:** The *gamification* of behavioral intervention for tic disorders (TDs) may not only enhance compliance with treatment protocols but also offer a key clinical advantage. By providing immediate positive feedback when tics are suppressed, games can counteract negative reinforcement processes that reinforce tics, which assumingly alleviates unpleasant premonitory urges. We developed a gamified protocol (XTics), which leverages this potential by combining gamified tic triggers with immediate reinforcement of tic suppression. We evaluate the clinical value of immediate reward contingency in enhancing tic suppression performance.

**Methods:** XTics comprises two conditions: tic-contingent and non-contingent. In the ticcontingent version, game progression was determined by real-time input from an experimenter who monitored the participant’s tics, rewarding tic suppression with favorable outcomes. Conversely, in the non-contingent version, game events occurred randomly. Using a crossover design, we trained 35 participants (aged 7-15) in both versions, with each participant undergoing a preliminary behavioral training and three hourly sessions for each condition. We both evaluated the overall protocol’s four-week impact on tic severity measures and compared contingent and non-contingent conditions.

**Results:** We achieved complete adherence to the protocol, while the participants increased their tic-free intervals by an average of 558% from the first to the last training day. YGTSS, a clinical measure of tic severity, showed an average clinically meaningful reduction of 25.69±23.39%, which was larger than that observed in control interventions and comparable to the effects of longer non-pharmacological treatments. Parent-reported tic severity decreased by 42.99±31.69% from baseline to three months post-treatment. When contrasting the ticcontingent with the tic-non-contingent training versions, we observed a larger improvement in tic-free interval duration in the former case (t(67)=3.15, p=.0025). Additionally, Rush, another measure of tic severity, demonstrated a greater reduction following training with the contingent compared to the non-contingent version (t(47)=3.47, p=.002).

**Conclusion:** The combination of gamified tic triggering with immediate and contingent rewards demonstrates a promising approach for enhancing treatment efficacy in TDs, offering an engaging boost to traditional therapeutic methods.

## Introduction

Tics, which are sudden, rapid, recurrent, and non-rhythmic involuntary vocalizations and/ or hyperkinetic movements^1^, typically emerge between the ages of 3 and 8 years^2^. Transient tic disorders (TDs) affects about 2.99% of the children, whereas the estimated prevalence of Tourette’s Syndrome (a persistent and multimodal form of tic disorder) is 0.77%^3^ in this population.

The etiology of TDs has yet to be elucidated. Various neuromodulators have been implicated in this disorder - including noradrenaline, glutamate, serotonin, opioids, acetylcholine, GammaAminoButyric Acid (GABA), and dopamine^4^ - but their combined contribution to its pathophysiology is still under research. However, as all first three drugs that were approved by the FDA for tic treatment have *dopaminergic* function^5^, and in light of a line of supporting evidence, dopaminergic abnormality, and particularly deficiency in dopaminergic neurotransmission in the cortico-basal-ganglia-thalamo-cortical loop, is widely considered as a major factor in TD. Maia and Conceição^6^ recently demonstrated how elevated levels of this neurotransmitter can facilitate prompt acquisition of tics via enhanced habit learning. In this process, the tic generates a sense of relief from the uneasiness of the premonitory urge. The relief is experienced as a reward, which is particularly powerful in the context of enhanced tonic and phasic dopamine levels that strengthen the direct relative to the indirect pathway.

Assuming that learning processes are at the root of TD, cognitive-behavioral protocols that specifically intervene with tic learning may be beneficial. Taking into account the adverse effects of drug treatment in TD^5,7–9^, non-pharmacological interventions offer considerable benefits, especially for children. Indeed, the advantage of behavioral therapy was recently confirmed by a meta-analysis, which found a medium to large effect size (0.67–0.94) in reducing TS symptoms^10^. A recent ^11^ workfound that behavioral therapies were as effective as pharmacological treatments (pimozide, aripiprazole or risperidone) in reducing symptoms and improving quality of life measures.

Accordingly, non-pharmacological therapies are recommended by current clinical guidelines for treatment in TDs^12–14^. These treatments include habit reversal therapy, comprehensive behavioral intervention (CBIT), and exposure and response prevention (ERP)^15,16^. Our work focuses on ERP, which was originally based on the assumption that tics are conditioned responses to premonitory urges. Hence, prolonged exposure to these urges without carrying out the conditioned responses (the tic) is expected to weaken the urge-tic association. Habituation to premonitory urges may follow, resulting in reduction in both urges and tics. ERP treatments usually start with two training sessions, in which the individual is taught to suppress all tics systematically. Suppression duration is timed, and individuals are motivated to beat their earlier records.

Behavioral treatments offer significant advantages for individuals with TD, but their effectiveness is hindered by several extrinsic factors. Firstly, there is a notable scarcity of clinicians trained in these methods^17^. A survey of American parents of children with TD revealed that 27.9% avoided behavioral treatment due to a lack of knowledge about where to find qualified clinicians, while 12.9% could not find a nearby clinician. Additionally, adherence to treatment protocols is challenging for many young patients, often due to comprehension difficulties or loss of interest^15,16^. An online survey found that approximately half of the parents reported their child’s behavioral treatment for TD was discontinued after five or fewer sessions, despite the recommended eight sessions over 10 weeks in CBIT and twelve in ERP for effective tic relief^18^.

To improve the accessibility of non-pharmacological interventions for TD, the development of computerized tools implementing CBIT or ERP components shows promise. Recently, English (TicHelper^19^, ORBIT^20^), German, and Swedish versions of such tools have been developed, with the web-based platform TicTrainer™ being particularly notable^21^. This tool, aligning with ERP principles, enables individuals to train in extending tic-free intervals while caregivers monitor and provide feedback on suppression success. Importantly, TicTrainer incorporates a *rewardenhanced* exposure strategy, making explicit incentives for users. This approach is underpinned by evidence suggesting that tic suppression is more effective when tic-free periods are rewarded, as opposed to noncontingent rewards (for review, see^22^), highlighting the significant influence of reward contingency on tic behavior. This phenomenon can be explained based on the notion that tics consolidate through negative reinforcement. In this process, performing a specific motor action or vocalization alleviates the aversive premonitory urge, thereby stabilizing them as tics^23^. Himle and colleagues^24^ suggest that rewarding tic suppression competes with the negative reinforcement provided by the tic, thus strengthening an alternative inhibitory response to the urge.

The present study introduces XTics, a gamified protocol designed to integrate treatment within a contemporary and widely favored medium among children. It is designed to augment ERP treatment in TDs, operable by co-therapist with minor training or even by caregivers. We conducted a six-session intensive training course for children with TD to evaluate its effects on tic suppression skills and daily tic severity. XTics aims to elaborate existing tic-treatment gamification. XTics, grounded in the rationale of immediate feedback’s importance in countering the tic’s negative reinforcement^24^, provides continuous and immediate feedback, both positive and negative. This approach aims to reinforce inhibitory processes and discourage tic behavior. XTics specifically focuses on boosting two pivotal aspects of TD intervention through gamification:

### Stimulation

Building on the concept that phasic dopaminergic activity is crucial in TDs and given that dopamine levels increase during reward anticipation (e.g.,^25,26^), we integrated multiple anticipation events in ZenithX to provoke tics. Our preliminary experiment^27^ showed that tic frequency indeed increases during these gamified expectation events. Consequently, unlike standard training protocols, ZenithX proactively engages users with various tic triggers. We also drew upon the idea that in ERP, exposing individuals to urges in various relevant contexts may not only boost engagement in treatment but also enhance tic suppression learning. Research indicates that varying the content, order of stimuli, and their context can boost exposure-based therapies^28,29^. Thus, XTics introduces a variety of tic triggering events.

### Reinforcement

In line with the evidence reviewed above, we expected that a game design that immediately rewards or penalizes its users depending on their success in elongating tic suppression will improve the training outcomes. We further adopted insights from the gaming research literature about the diverse reward mechanisms in video games. Philips and colleagues^30,31^ identify six types of video game rewards: *glory*, *sensory feedback*, *facility*, *sustenance*, *access*, and *praises*. Current training protocols, including TicTrainer, primarily utilize glory rewards, which emphasize user achievements without affecting real-time gameplay. Our game, ZenithX incorporates a broader spectrum of reward mechanisms. These include *facility* rewards that enhance player capabilities (e.g., upgraded equipment), *sustenance* rewards to counteract negative gameplay states (e.g., health restoration), and *sensory feedback* for an enriched aesthetic experience, thereby diversifying the motivational elements in the training process.

The expansion and diversification of immediate rewards in our approach aims not only to enhance user engagement but also to cater to the diverse needs and reward sensitivities of different users. For instance, unmedicated individuals with TS tend to learn more effectively from gains rather than losses, a trend that reverses in individuals on antipsychotic medication for TS (see ^6^). We expected that a varied range of gamified rewards that adapt to such differences in learning and motivational responses will enhance training effectiveness.

Thus, our group developed ZenithX (Figure 1a-b, Video S1), which combines elements from the card game *War* and the board game *Ladders and Snakes*. In this video game, the participants compete with bots as they move their tokens along a numbered grid, aiming to be the first to reach the end of the route. The players may engage in card-battles where they play and later regain cards (i.e., *facility* and *sustenance*). The strength of the bot opponents and the expectancy of the card regained from an animated chest are represented by the colors of the token and chest (Gold > Silver > Wood).

**Figure 1:**
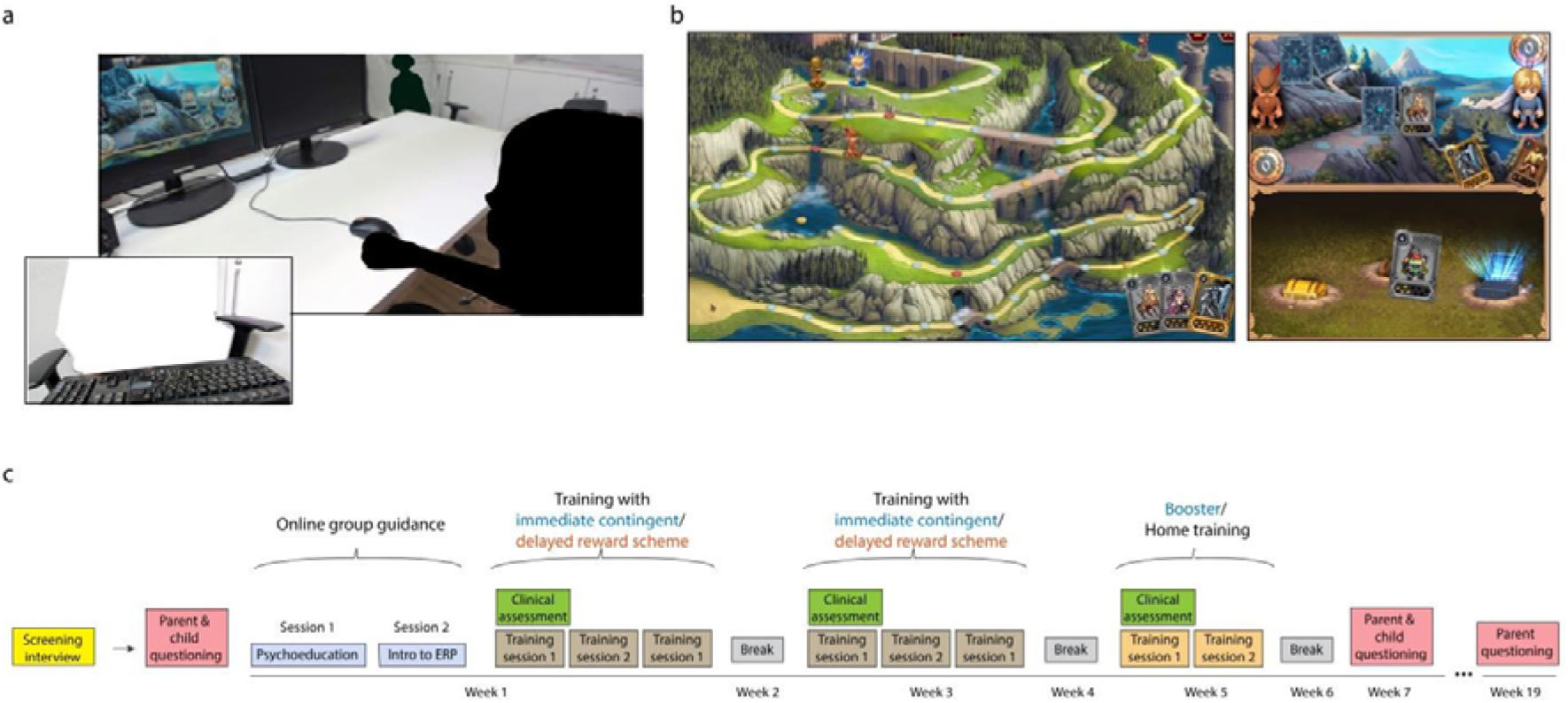
Closed-loop rationale and the outline of XTics. (a) A participant engages in playing *ZenithX*, while the experimenter actively monitors for tics, pressing a key upon detection. The image serves an illustrative purpose; No participant was involved in the production of this photograph. (b) The participant’s success in suppressing tics influences their probability of succeeding in specific game events including dice rolls, battles, and card regaining. (c) The study outline.

Importantly, ZenithX allows real-time experimenter input that influences the gameplay events of dice rolls (DICE), battles (BATTLE), and card regain (CHEST). The experimenter marks a tic occurrences, impacting game dynamics. To understand how this immediate, detailed feedback assists participants in managing tics, we used a crossover design (Figure 1c), implementing two conditions. In the Immediate and Contingent Reward (ICR) condition, tic suppression directly affects gameplay, with participants informed that suppressing tics weakens opponents and improves card acquisition. Daily sessions end with feedback on tic suppression performance relative to that day’s baseline. Conversely, the Delayed Reward (DR) condition’s gameplay does not depend on performance, but participants still receive end-of-session feedback on tic suppression. We anticipated enhanced tic suppression performance following the DR condition, which incorporates the previously mentioned stimulation principles. Furthermore, we expected even more substantial improvement after the ICR condition due to the integration of both stimulation and reinforcement elements.

In line with the ERP rationale, our primary measure of tic suppression was tic-to-tic interval (TTTI); i.e., the duration of an interval between two successive tics. Our hypotheses are specified in Table 1. We registered the study before analysis at the Israeli Ministry of Health clinical trails website (https://my.health.gov.il/CliniTrials/Pages/MOH_2023-05-02_012595.aspx) and the Open Science Framework (https://doi.org/10.17605/OSF.IO/AF74W; See Supplementary Materials for modifications to the analysis plan). We compared XTics outcomes with those of control and alternative tic treatments using data from a study involving two similar age groups receiving either CBIT or psychoeducation (H11-15). Finally, half of the participants completed two additional daily booster sessions with ICR (H17-18).

**Table 1:**
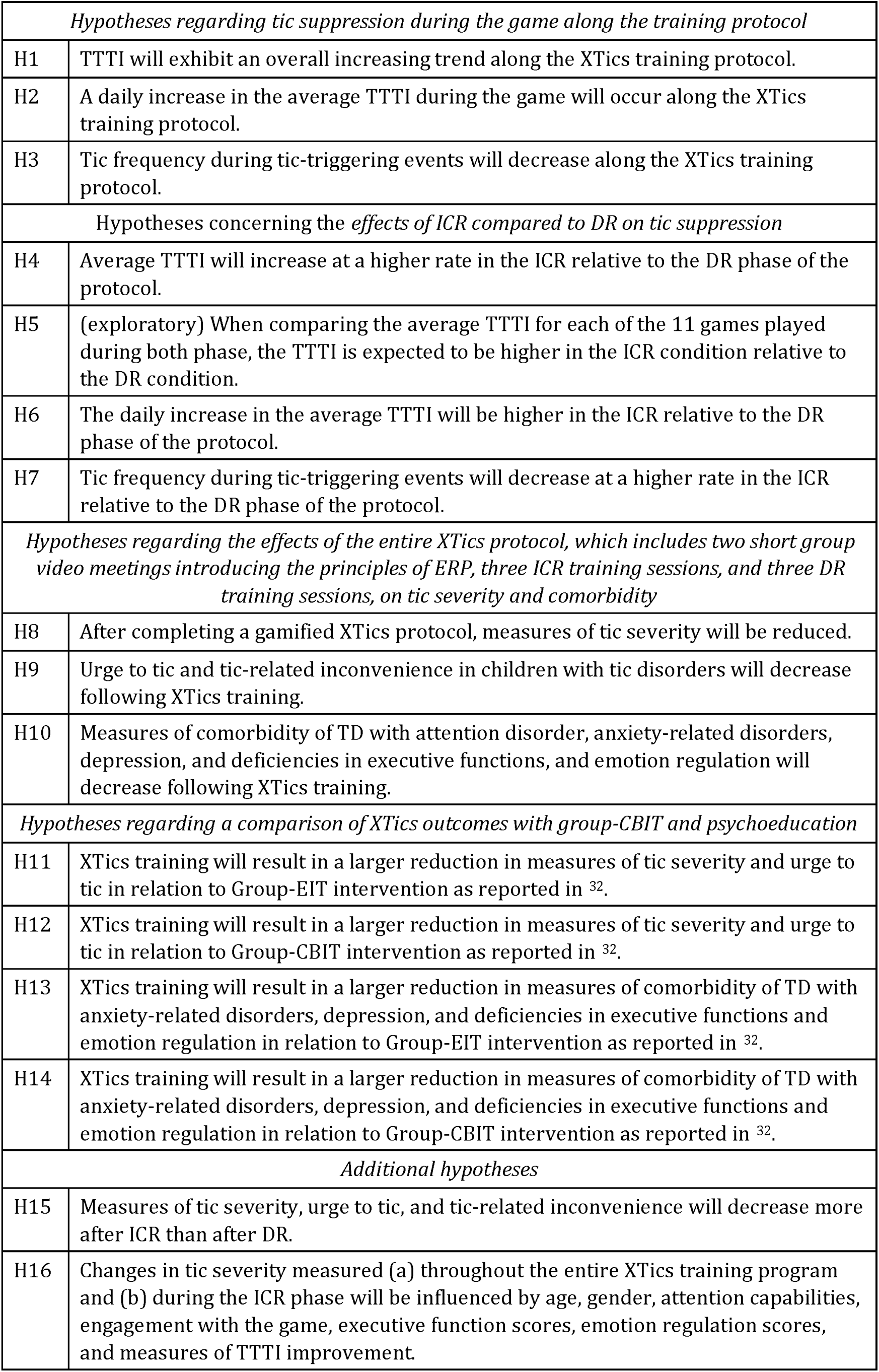
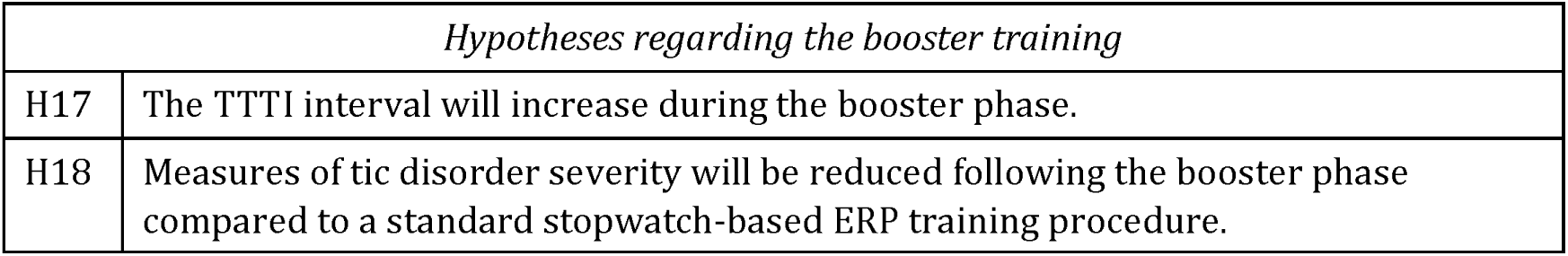
Research hypotheses.

## Methods

### Participants

The participants were recruited between July and September 2022 through personal contacts or advertisements published at the Pediatric Movement Disorders Clinic at the Pediatric Neurology Unit, Tel Aviv Sourasky Medical Center, and the Tourette Syndrome Association in Israel. Out of 102 respondents, 50 children and adolescents underwent screening interviews, and 38 were accepted for treatment.

During the screening interview, the clinician and parent conducted a simultaneous tic frequency count while the child watched three short animations^27^. Following the screening, a brief Hebrew version of the Anxiety Disorders Interview Schedule for Children (ADIS-C/P)^33^ was administered to the parent to assess comorbidities. Parents and children were then invited to complete online questionnaires designed to further assess comorbid symptoms. Written informed consent to participate in the full study was obtained from parents at the beginning of the first ERP personal tutorial and clinical assessment session at the Sagol Brain Institute, TelAviv Sourasky Medical Center.

Inclusion criteria were: (1) children and adolescents aged 7–15 years, (2) with at least moderate tic severity as indicated by a Yale Global Tic Severity Scale (YGTSS) tic severity score≥11 and (3) Tic frequency of over one tic per minute on average in the screening interview. Exclusion criteria were:(1) behavioral treatment for tics in the past 12 months, (2) pharmacological treatment for tics that has not been stable for the past 6 weeks or with planned changes during study participation, (3) evidence of tics that may produce physical harm to the child and (4) a history of psychiatric or neurological disorders requiring hospitalization or a known cognitive decline. Since TS is seldom seen without comorbidities^34^, co-occurring anxiety disorders (n=7), ADHD (n=12) and learning disabilities (n=1) were included, unless the disorder required immediate treatment or change in current treatment. Eight patients were taking medications (amphetamines=6; clonidine=2; SSRIs=1), and eight went through a behavioral intervention for TD in the past (CBT=6; CBIT=1; ERP=1).

Three participants that passed the online screening were excluded: (i) a participant that scored only 8 at the tic severity scale of the YGTSS at the clinics; (ii) a participants who scored 10 at the tic severity scale, but manifested no tics during the first baseline game and asked to quit the experiment; (iii) a participant who over-reacted violently to game losses and did not follow the instructions so that his tics were not properly coded. Another participant who passed the online interview and scored 10 in the clinical assessment of tic severity was nevertheless accepted, since he manifested a considerable number of tics in the baseline game. One participant was excluded due to repeated lateness. The final cohort consisted of 35 patients (12 girls; age: 10.31±2.61, range: 7-15 years).

### Materials

#### The game (Figure 1, Video S1)

In ZenithX participants compete with three virtual opponents in a race to the top. The opponents differ in color, which indicates their strength (their success rates in battles). A virtual dice dictates the number of forward steps for each of the players in each round. When two players occupy the same slot, a combat is initiated. The combat includes the following phases: (i) *Card selection* : the participant selects one out of three cards at her hand. The cards are numbered from 1 to 9 where 9 is the strongest card. (ii) *Battle*: The selected card competes with the opponent’s card. The winner moves forward, and the loser retreats a number of steps that equals the difference between the competing cards’ numbers. In case of a draw, the battle restarts with the remaining cards; (iii) *Card retrieval* : An animation of three chests appears on the screen. The chests differ in color, which indicates the strength of the cards they contain (i.e., the chests differ in card’s number probabilities). One of the chests is shaking during an anticipation interval. Finally, the chest opens, and a card is exposed and retrieved by the participant.

In line with the ERP rationale, the participant is encouraged to avoid tics over increasing periods of time. In the ICR phase of the experiment, we calculated a baseline average TTTI (ATTTI) (see below). Next, we computed the average gap (AG) between successive TTTIs that are equal or higher than the ATTTI. During the game, the participant’s tic manifestation affects four core game events: (i) dice outcomes (higher scores for longer TTTIs), (ii) opponent’s card (weaker cards for longer TTTIs), (iii) chest type (favorable chests for longer TTTIs), and (iv) retrieved card (stronger cards for longer TTTIs). For each of these four elements, we define seven preset probability distributions, with a gradient of chance to obtain desired outcomes. For example, the preset probabilities to obtain wood, silver, or gold chests range between [0%,0%,100%], [0,20%,80%], [0%,50%,50%,], [26%,48%,26%], [50%,50%,0%], [80%, 20%,0%], and [100%,0%,0%], respectively. Similarly, the chance of retrieving a strong card is higher in the case of gold relative to other chests.

Depending on the participant’s TTTIs in the ongoing game and on the baseline ATTTI and AG, a preset is selected so that the chance to obtain a desired outcome improves with longer TTTIs. For each core game event, a composite performance score is calculated by generating a vector of the participant’s TTTIs from the game’s start until that event, counting only TTTIs that are equal to or larger than the ATTTI. The interval between the last registered tic and the current core game event is computed as TTTI as well. The timing differences between the core game event and the TTTIs endings are calculated, and weighted by an inverse sigmoid function *f*. The composite measure for a certain core game event, corresponding to the *j*th TTI in the game, is then computed as follows:

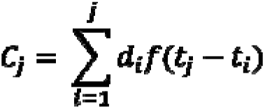

where ***d_i_*** is the duration of a ***TTTI_i_***, and ***t_i_*** is the timing of the TTTI ending. The inflection point of *f* depends on the baseline ATTTI so that in steeper sigmoid for shorter ATTTIs, TTTIs that ended long before the current core game event have only negligible effect on the composite score, and vice *versa*. The inflection point’s maximal value was set to 25 seconds. The composite measure ***C_j_*** is then divided by the simulated ***Cs***, which is computed using the same formula, but with a synthetic vector of TTTIs identical to ATTTI, separated by AG. Thus, the event score 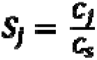 represents the participant’s improvement relative to the baseline at the specific point in the game. According to predefined ***S_j_*** ranges, one of the seven presets is selected so that high ***S_j_*** values yield distributions with higher chance for favorable outcomes.

Battle card presets are defined for each opponent so that the Gold bot is stronger than the Silver, which is stronger than the Wood bot. When battling against the participant, the battle card presets are selected according to ***S_j_***, as described above. In the randomized battles between bots, other presets that maintain the power relations between bots are used.

The game ends when one of the players reaches the final slot. The game score is calculated as follows: each player earns 100 points minus their distance from the final slot multiplied by 5. The first and second players receive a bonus of 10 and 5 points, respectively.

Each of the ICR and DR phases included three training days, with the first round instructing participants not to suppress tics and providing the baseline parameter for the first day. The baseline for days 2 and 3 is computed based on data from all training runs on days 1 and 2, respectively. Under both conditions, participants were instructed to avoid tics while continuously practicing the learned ERP principles during the game.

### Intervention protocol

The training protocol (Figure 1c) included a preliminary psychoeducation tutorial, one week of ERP exercise under the ICR condition, a week off treatment, a week of exercise with the DR version of the game, a second week off treatment, and a booster/treatment-as-usual week. The order of the test and control exercise weeks was randomized and counterbalanced.

The protocol was administered by a research team that included clinicians and co-therapists (see Supplementary Materials). Introduction to ERP and psychoeducation was delivered to all participants in two 90 minutes virtual group sessions (via ZOOM) conducted by a senior cognitive-behavioral therapist (SZB) with expertise in this treatment The virtual group sessions were performed separately to participants of younger (7-10 years) and older (11-15 years) age. These sessions focused on psychoeducation on tics and ERP principles and also included the acquisition of relaxation techniques such as deep diaphragmic breathing and progressive muscle relaxation. Following the group sessions, the child and parent met with a clinician to monitor and support the course of ERP exercises. Tic severity assessment (YGTSS, PUTS, PTQ and Rush) was administered at each of these sessions by an independent trained clinician who was blinded to phase assignments (test/control).

ERP exercise sessions were held individually and conducted by a co-therapist. Each child attended three exercise sessions per week for each of the two phases. During the first exercise session, children were introduced to the game rules via a short presentation delivered by the co-therapist, followed by a short trial game. Children were informed that they would play successive game rounds each day, with scores accumulated between days for each player. The highest accumulated scores displayed on the scoreboard by the end of the last exercise session would determine the winners, who would receive a prize of 100 ILS.

At the first weekly exercise, the child played a baseline game while instructed to “feel free to tic,” as the co-therapist counted the tics. The co-therapist was positioned in front of the child (Figure 1a), registering tics by pressing the space-key. After completing the baseline round, further instructions were given, depending on the child’s phase assignment. The co-therapist then operated the exercise rounds, encouraging the child to avoid tics by applying the learned relaxation techniques.

On the first day, the participants played a baseline game and three training games, while on days 2 and 3 they played four games. Typically, a game lasted around 12 minutes, while the entire exercise session was 60 minutes long. At the end of each training week, the co-therapist asked the participants to try to identify the contexts and/or factors that exacerbate their tics (e.g., social engagement, watching television). The participants were then asked to practice relaxation techniques during these situations when they happen the following week, and each of these practice sessions was counted by the research team as equivalent to a win in a game round (which would bring them closer to the prize). Parents were informed that the child was assigned this task for the following week and were asked to encourage him/her to complete it.

In Week 5, participants were randomly assigned to a Booster or Home-training group. The Booster group attended two additional exercise gamified sessions per week, while the Hometraining group followed the standard stopwatch-based ERP procedure, explained by the clinician, for two hours per week. Upon completing the training, participants received a printed letter of appreciation and a monetary prize.

#### Measures

All measures are specified in Table 2. Tic severity was assessed by an independently trained clinician, the co-therapist, parents, the children themselves, and through objective counting. Additionally, we evaluated tic suppression performance during gameplay: Tics were identified and recorded in real-time by a co-therapist who continuously monitored the participant while playing. If the participant exhibited a sequence of tics, the co-therapist recorded approximately one tic per second.

**Table 2:**
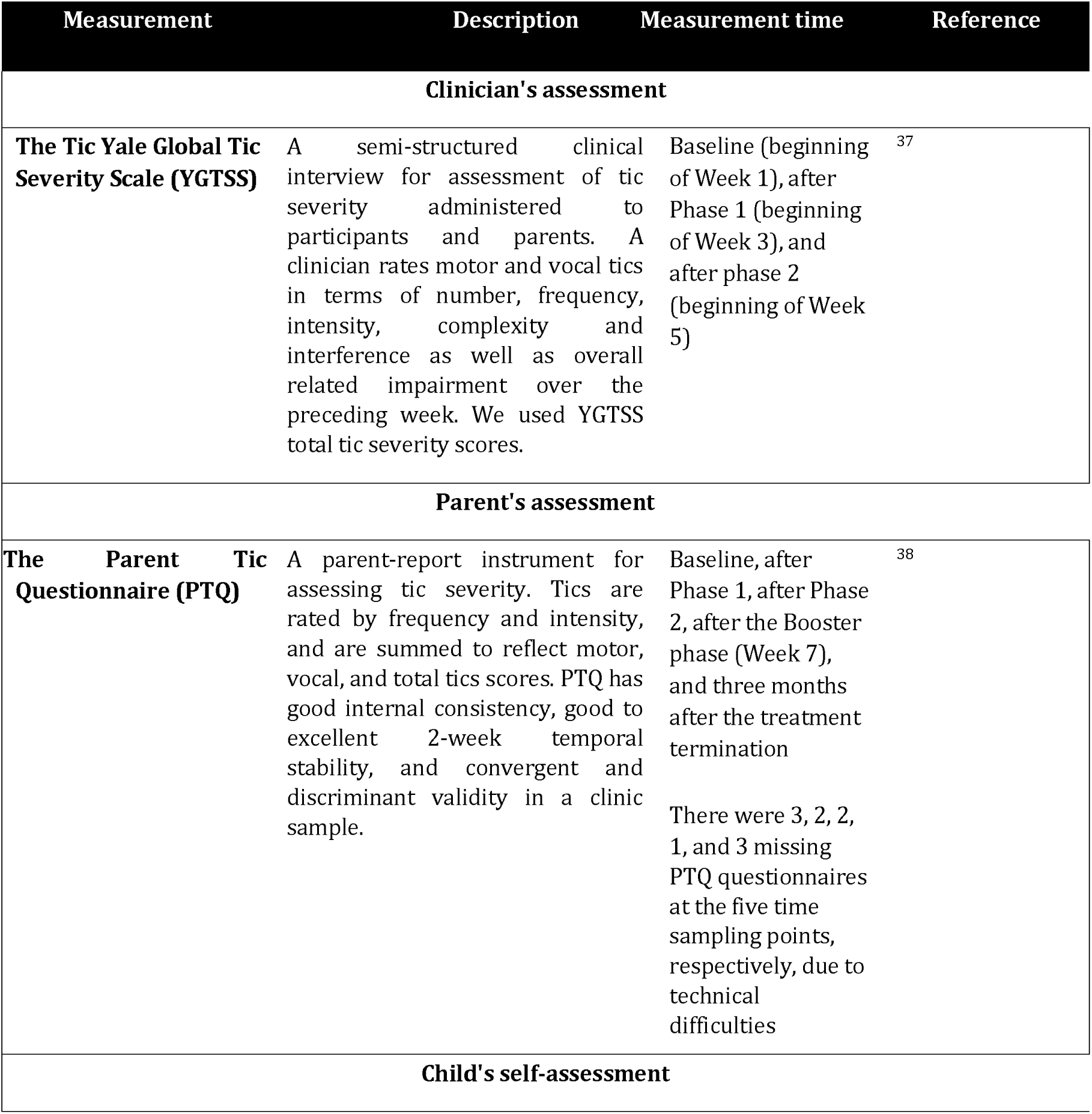
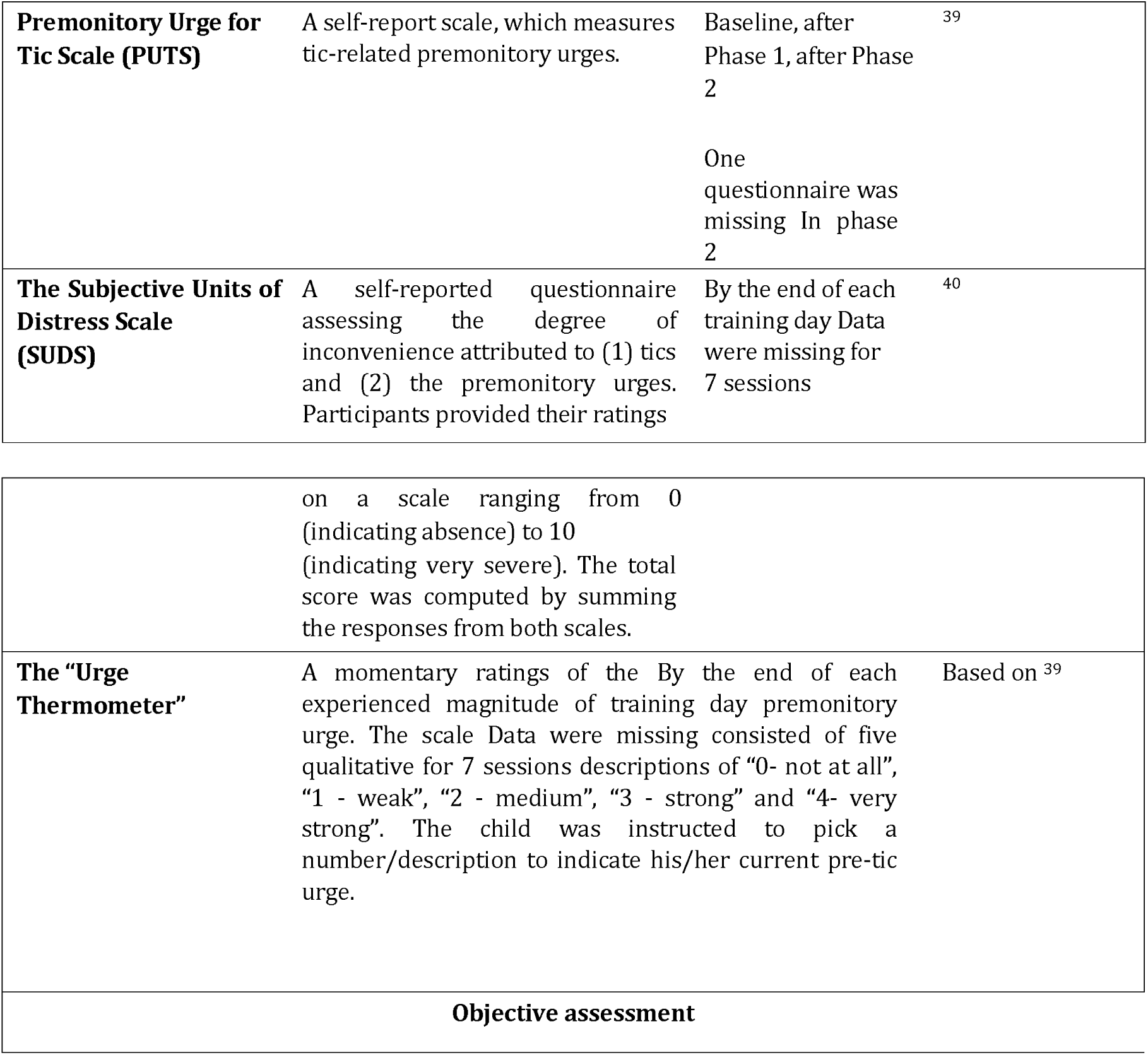
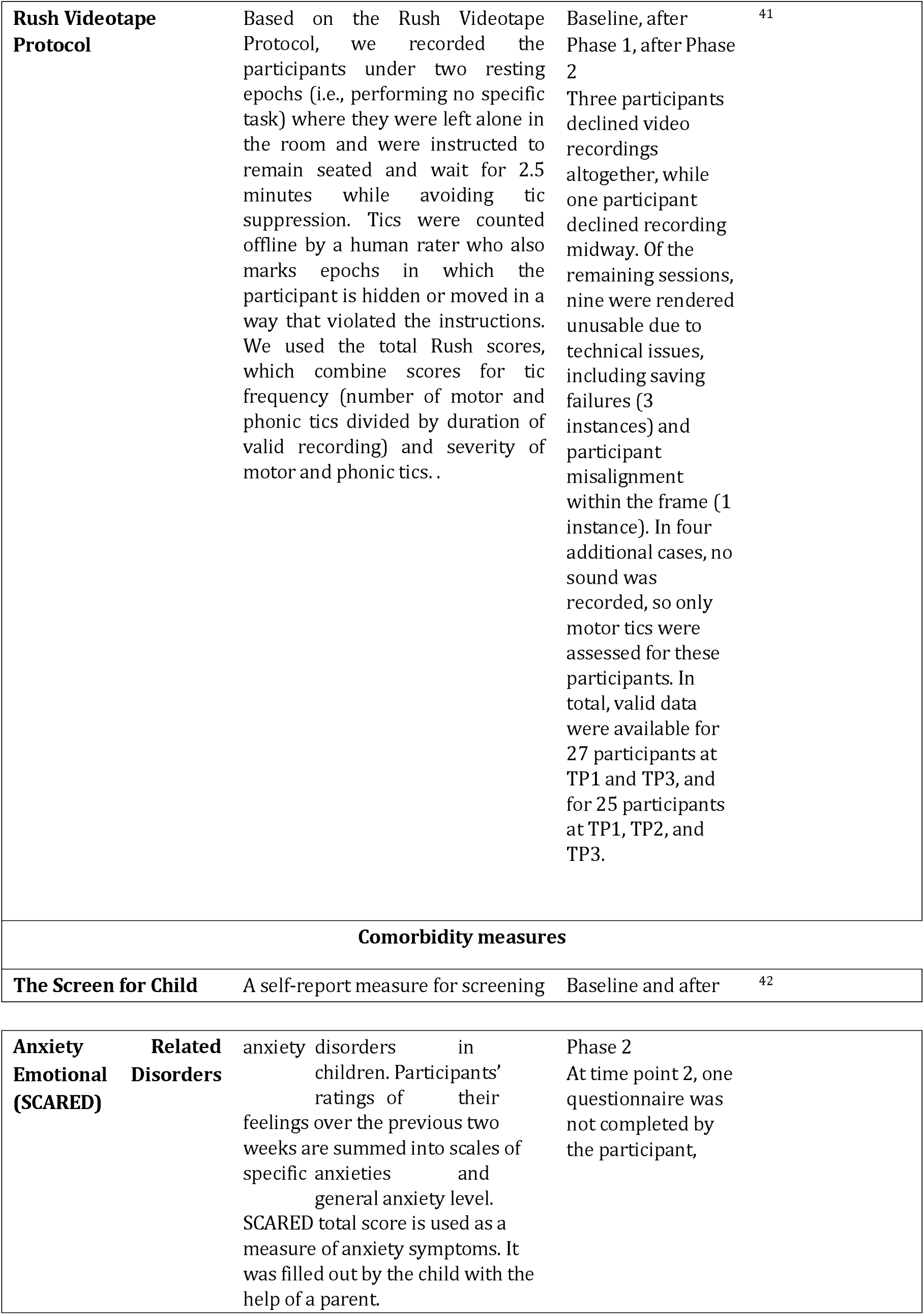
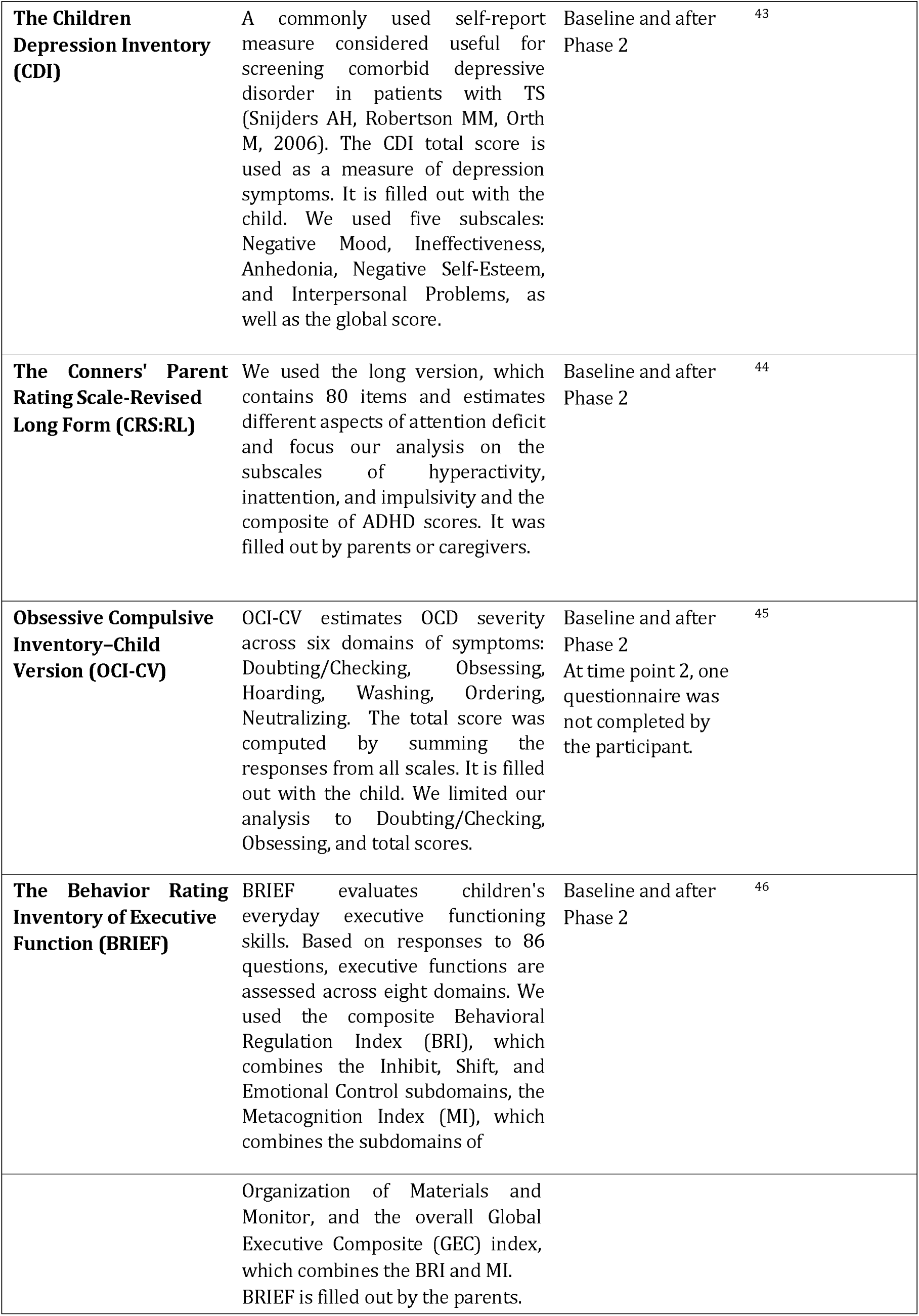
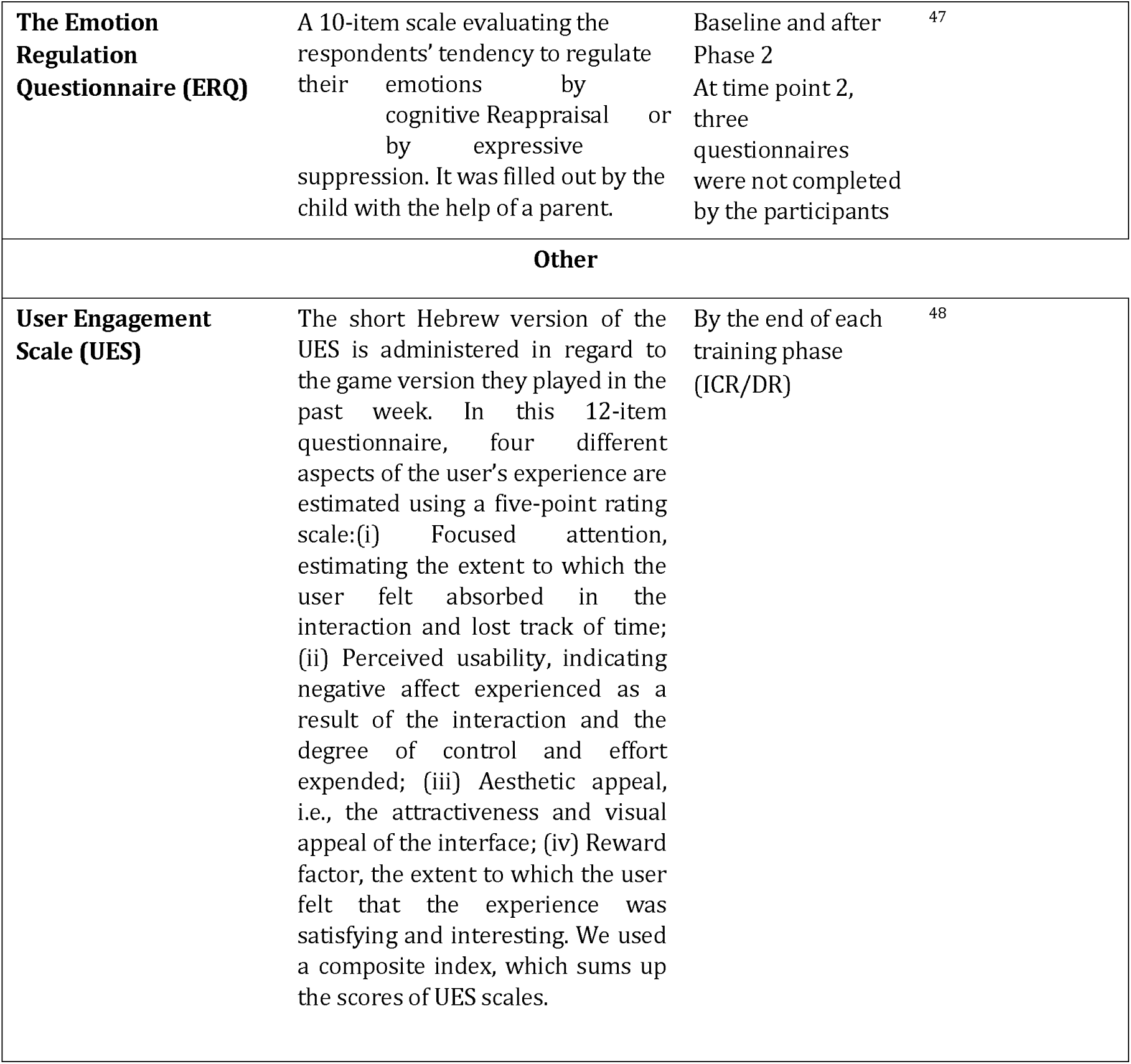
Psychiatric and psychological measures employed in the study.

To assess the reliability of tic coding, we compared online recordings with offline annotations conducted by independent raters for the same seven randomly selected sessions. Tics were resampled in one-second intervals, and inter-rater reliability was assessed using Cohen’s Kappa index^35^. Apart from one session where the agreement level was fair (Cohen’s Kappa of 0.27), all other sessions achieved moderate to substantial agreement (0.53-0.8), indicating a generally reliable annotation process^36^.

### Analysis

H1: To estimate the change in TTTI throughout the XTics protocol, we z-tested Fisher-z transformed Pearson coefficients for the correlation between the TTTI duration and its ending time along the time axis of the entire protocol.

H2: To estimate the daily change in TTTI throughout the XTics protocol, we averaged the TTTI for each of the six days and z-tested the Fisher-z transformed Pearson coefficients for correlation between the daily average TTTI duration and its ending time.

H3: To assess the impact of training on tic frequency during DICE, BATTLE, and CHEST events, we examined changes in tic frequency during all three events separately throughout the XTics protocol by z-testing the Fisher-z-transformed Pearson coefficients, measuring the correlation between tic frequency in each event and its ending time along the time axis of the entire protocol. Additionally, to estimate the change in daily mean tic frequency during tic-triggering events, we calculated the average frequencies for each of the six days. Then, we z-tested the Fisher-z-transformed Pearson coefficients to measure the correlation between the average frequency and time.

H4: The difference between the training conditions in TTTI was examined via a linear mixed model with two factors: (1) CONDITION: ICR and DR; (2) ORDER: assignment of ICR\DR to the first\second phase. The dependent variables were Fisher-z-transformed Pearson coefficients for the correlation between TTTI duration and their ending time within the tested experimental phase. The model incorporated the testing of intercepts. An additional exploratory dependent variable was area under the curve (AUC) quantifying the cumulative TTTI change across sessions, calculated using the trapezoidal rule and normalized by the mean TTTI and number of samples for each individual.

H5: In each of the two phases, average TTTI in the baseline game and each of the 11 training games was computed. For each training game *g* we calculated the difference from the baseline *D* as follows: 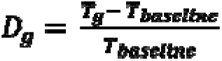 where *T* is the average TTTI. In one case, one of the games was discarded as the participant quit it. *D* values were then compared between phases for each of the games using Wilcoxon signed rank test. To control for multiple comparisons, we employed the FDR correction.

H6: Daily change in TTTI was compared between training conditions via a linear mixed model with two factors: (1) CONDITION: ICR and DR; (2) ORDER: assignment of ICR\DR to the first\second phase. The dependent variables were: (a) Fisher-z-transform coefficients for a three point correlation between the average daily TTTI and the day number; (b) 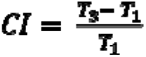 where ***T*_1_** and ***T*_3_** are the average TTTI on the first and last day of the phase, respectively.

H7: We computed tic rate during tic-triggering events of DICE, BATTLE, and CHEST considering either all three event types or only the two latter types. The hypothesis was tested using a linear mixed model with two factors: (1) CONDITION: ICR and DR; (2) ORDER: assignment of ICR\DR to the first\second phase. We employed three alternative dependent measures: (i) Fisher-ztransformed coefficients for a correlation between the tic rate in each of the events and the aggregated ending time of that event. (ii) Fisher-z-transformed coefficients for a three point correlation between the average daily tic rate during the tic-triggering events and the day number; (iii) 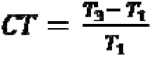 where ***T*_1_** is the average tic frequency in the first day of the phase and ***T*_3_** is the average tic frequency in the third and last day of the phase.

H8: Decrease in tic severity was assessed via one-sided Wilcoxon tests comparing post-pre XTics difference in YGTSS (total tic severity), PTQ (total tic severity), and Rush scores.

H9: Given that the ERP treatment is designed to heighten a child’s awareness of the urge-to-tic sensation and its associated inconvenience, we hypothesized that these parameters might exhibit an initial increase followed by a subsequent decrease over the course of the treatment. Consequently, two alternative analyses were conducted: (1) A one-sided z-test of the Fisher-ztransformed Pearson coefficients for the correlation between SUDS or urge thermometer scores and the sample time. Additionally, we tested for post-pre phase difference in the case of PUTS scores as in H8; (2) The difference between the final scores and the maximum SUDS and urge thermometer scores recorded on day 1 and day 2 was computed. A one-sided t-test was subsequently conducted on these score differences. The resulting t-statistics were then compared to a null distribution derived from 100,000 analogous tests with time labels shuffled randomly.

H10: Decrease in comorbidity scores following the treatment was assessed using one-sided Wilcoxon tests comparing post-pre XTics difference in SCARED, CDI, CRS:RL, OCI-CV, BRIEF, and ERQ scores.

We compared XTics outcomes with psychoeducation and CBIT outcomes for a similar age group (8-15 years old) with YGTSS severity scores of 14 or higher^32^. The study included eight weekly sessions structured similarly to ours (two 90-minute sessions followed by six 60-minute sessions). The participants were divided into two groups: Group-CBIT (n=27, average age=11.11±1.67) received CBIT, while the control group (Group-EIT; n=28, age: 10.74±1.24) received only educational intervention for tics with no training.

H11: To contrast reduction in tic severity and urge measures with the group-Education Intervention for Tics (group-EIT), we performed one sided Mann-Whitney tests, comparing post-pre XTics of YGTSS, PTQ and PUTS score differences between the XTics and EIT groups.

H12: To contrast reduction in tic severity measures with the group-Education Intervention for Tics (group-CBIT), we performed a one sided Mann-Whitney tests, comparing post-pre XTics YGTSS, PTQ and PUTS score differences between the XTics and CBIT groups.

H13: To contrast reduction in comorbidity measures with the group-EIT, we performed onesided Mann-Whitney tests, comparing post-pre XTics SCARED, CDI, BRIEF, and ERQ score differences between the XTics and EIT groups.

H14: To contrast reduction in comorbidity measures to the group-Education Intervention for Tics (group-CBIT), we performed a one-sided Mann-Whitney tests, comparing post-pre XTics SCARED, CDI, and ERQ score differences between the XTics and CBIT groups.

H15: To compare reductions in tic severity measures across training conditions, we utilized a linear mixed model incorporating two factors: (1) CONDITION, encompassing ICR and DR; and (2) ORDER, representing the assignment of ICR or DR to the initial or subsequent phase. Our dependent variables comprised post-pre differences in YGTSS, PTQ, and Rush total scores for each respective phase. Premonitory urge and tic-related distress measures were tested via the same linear mixed model where two alternative SUDS and urge thermometer measures were used: (1) Fisher-z scored Pearson coefficients for the correlation between SUDS/urge thermometer scores and the sample time; (2) The difference between the final scores and the maximal scoring in day 1 and day 2 of each phase.

We assessed the statistical significance of the main effects of CONDITION and ORDER by comparing their corresponding t statistics to null distributions generated through 100,000 permutations, wherein either condition or time labels were randomly shuffled. CONDITION×ORDER interaction was similarly tested against a null distribution resulting from the shuffling of both time and group labels.

H16 (exploratory): We employed Spearman correlation analysis to explore associations between post-pre X-Tics and ICR phase difference in YGTSS, PTQ, and Rush scores on the one hand, and age, Conners’ ADHD scores, UES, ERQ total scores, Brief scores, and measures indicating changes in TTTI change. These measures include Pearson coefficients for the correlation between the TTTI duration and its respective ending time, 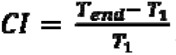 where ***T*_1_** and ***T_end_*** are the average TTTI on the first and last day of the phase, and TTTI AUC calculated as in H4. To explore gender-related differences, changes in tic severity measures between male and female participants were compared utilizing the Wilcoxon rank sum test. FDR correction was applied for all post-pre XTics and ICR phase tests separately.

H17: To test increase in TTTI during the booster training, we computed Fisher z-transformed Pearson coefficients for the correlation between tic-to-tic interval duration and their ending time within this experimental phase. We then conducted a one-sided z-test on the resulting values to estimate the significance of the results.

H18: The impact of the booster training was assessed using a Mann-Whitney test, comparing the difference between the PTQ scores before the booster/treatment-as-usual phase and the followup scoring (three months later).

## Results

### Adherence

None of the participants who entered the study dropped out voluntarily before the completion of two weeks of training. In one case, a child refused to attend two additional booster sessions. In another case, one participant was excluded from the study based on the researcher’s decision due to repeated lateness. The user engagement scores were 47.59±6.57 and 46.11±9.53 out of 60 in the first and the second phase, respectively.

### Alterations in tic-to-tic interval and triggered tics

To estimate the change in TTTI throughout the XTics training (H1,H2), we z-tested Fisher-ztransformed Pearson coefficients for the correlation between TTTI duration and the ending time of this interval along the time axis of the entire protocol. Regardless of the order of phase types (ICR/ DR), we observed a strong positive TTTI increase throughout the experiment (^-^=0.2±0.22, z=5.03, p<5×10^-7^, two-sided test). The daily TTTI average increased as well (^-^=0.49±0.41, z=6.73, p<1×10^-11^).

Accordingly, we observed a gradual decrease throughout XTics training in tic frequency during the three tic-triggering events (H3): DICE (^-^=-0.13±0.21, z=-3.54, p=.0005), BATTLE (^-^=-0.22± 0.26, z=-4.87, p<5×10^-6^), and CHEST (^-^=-0.22±0.28, z=-4.7, p<5×10^-6^). This trend was clearly evident also when examining changes in the average daily tic frequency: DICE (^-^=-0.4±0.35, z=6.01, p<5×10^-9^), BATTLE (^-^=-0.4±0.36, z=-6.16, p<1× 10^-9^), and CHEST (^-^=-0.51±0.31, z=-7.57, p<1×10^-14^).

Notably, at the group level TTTI increased from Day 1 to Day 6 by 678% and 235.49% on average and median, respectively (Cohen’s d=0.57). The average individual TTTI increased by 558% between the first and last training day, with a median increase of 135%. Tic frequency during the anticipatory events of DICE, BATTLE, and CHEST also considerably decreased from Day 1 to Day 6 in 59.1%/32.1% (mean/median)(d = 0.76), 77.9%/50.25% (d=0.85), and 74.4%/45.58% (d=0.96), respectively.

Figure 2a illustrates increase in TTTI throughout the experiment of a representative participant. The higher slope of the regression line in the ICR phase suggests that this condition facilitated efficient learning of the tic suppression technique. To systematically estimate the impact of the feedback type on training success, we compared coefficients of TTTI-time correlation between the ICR and DR conditions using mixed linear model (H4). We found that feedback CONDITION significantly affected TTTI change rate with an estimate of 0.2±0.065, t(67)=3.15, p=.0025, Q_FDR_<.05. The predictor ORDER also significantly affected the dependent variable with a coefficient estimate of 0.18±0.07, t(67)=2.41, p=.02. As demonstrated in Figure 2b, TTTI expansion was faster under immediate and contingent relative to delayed feedback, and this phenomenon was stronger among participants who trained with DF first and ICR last. However, the CONDITION×ORDER interaction was insignificant (t(66)=0.97, p=.34).

**Figure 2:**
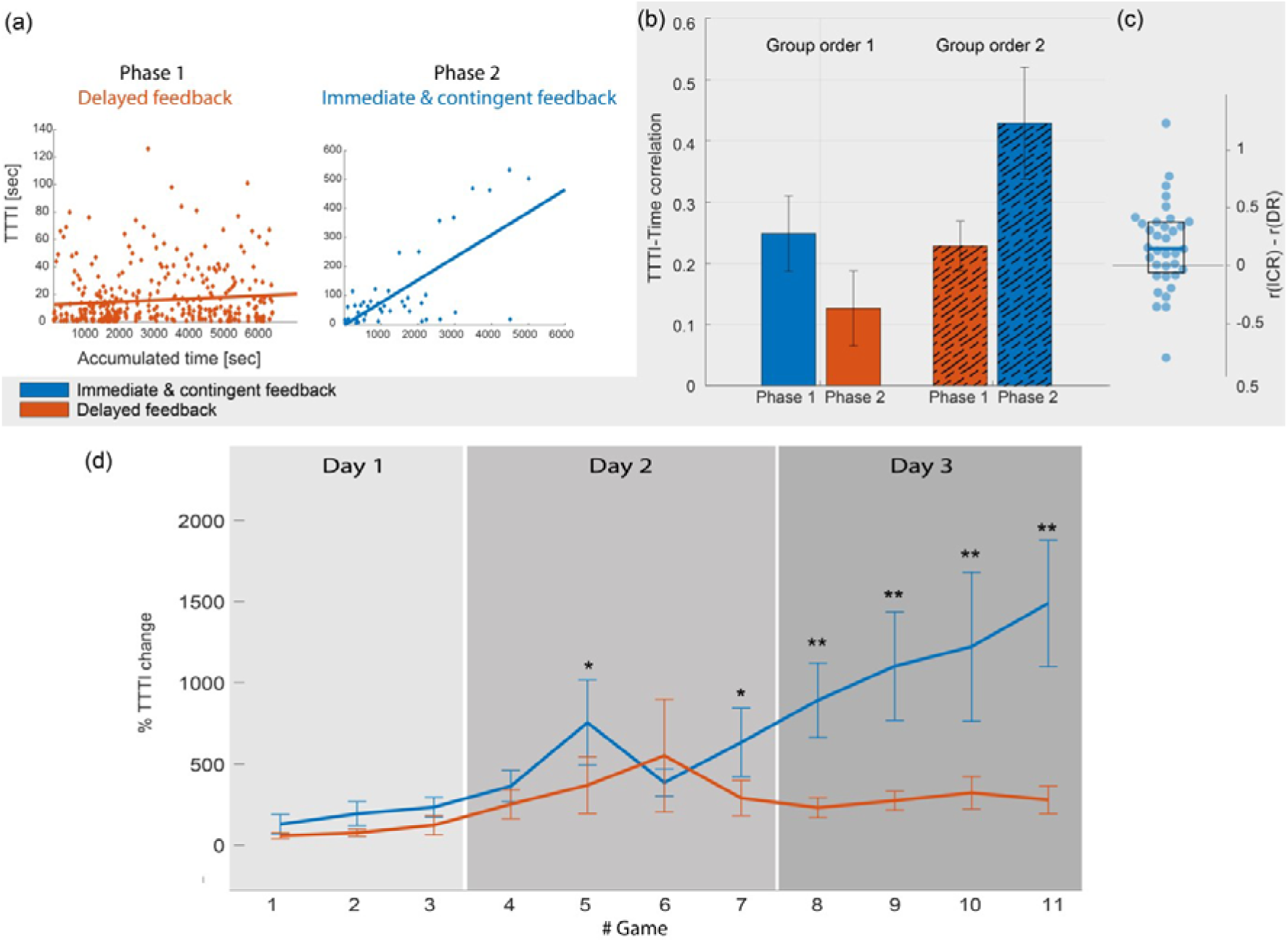
Tic-to-tic interval change in the test and control training conditions. (a) Data of a representative participant. Each dot represents a single TTTI. Regression lines are presented. The steeper slope of the regression line in the ICR phase suggests that this condition facilitated more effective of the tic suppression technique. (b) Averages and standard errors of Pearson coefficients for the correlation between TTTI duration and TTTI ending time during the two training phases under test and control conditions. This figure has been designed using assets from Freepik.com. (c) Differences between the Pearson coefficients for the ICR and DR conditions per participant. The central horizontal line signifies the median, while the upper and lower horizontal lines correspond to the 75th and 25th percentiles, respectively. (d) Average TTTI and standard error per game throughout the training phase for ICR and DR conditions. N=36, * Q_FDR_<.05, ** Q_FDR_<.005.

A comparative analysis of TTTI change over time (H5, Figure 2d) reveals an intriguing temporal pattern. TTTI change from baseline was significantly higher in ICR compared to the DR phase only in the fifth and the last five games (Z=2.34, p=.02, Q_FDR_=.04; Z=2.64, p=.008, Q_FDR_=.02; Z=3.51, p=.0005, Q_FDR_=.003; Z=3.1, p=.0009, Q_FDR_=.005; Z=3.31, p=.0006, Q_FDR_=.003; Z=4.11, p=5×10^-5^, Q_FDR_=.00004, respectively).

Similar to H4, a fixed effect of CONDITION (i.e., higher TTTI under ICR) did not survive FDR correction when using an alternative analysis of comparing TTTI percent signal change between the first and the last training days (t(67)=2.12, p=.03; H6). Neither significant effect of ORDER (t(67)=0.06, p=.95) nor CONDITION×ORDER interaction (t(66)=0.67, p=.5) was found in this case. We found significant effect for neither of these factors when examining the correlation coefficients for average TTTI by day number (CONDITION: t(67)=1.6, p=.12; ORDER: t(67)=1.21, p=.23, CONDITION×ORDER: t(66)=0.79, p=.43). When examining a third measure of tic suppression – TTTI area under the curve (AUC), we observed a main CONDITION effect (t(66)=3.85, p=.0003, Q_FDR<_.05) and CONDITION×ORDER interaction (t(66)=3.85, p=.003, Q_FDR_<.05), but no ORDER effect (t(66)=1.96, p=.06). This indicates that the normalized TTTIAUC was higher under the ICR condition, particularly when this condition was introduced during the second week of training.

In general, the enhanced tic suppression during ICR was not limited to the validated tictriggering events. We found no effect of CONDITION on tic rate during tic-triggering events both in a model that examines the tic rate in DICE, BATTLE, and CHEST events (t(67)=0.96, p=.33) and in a model that regards only the two latter event types (t(67)=0.7, p=.48). The latter model pointed to an effect of ORDER (t(67)=2.17, p=.03), but it did not survive correction for multiple comparisons. The ORDER effect was not significant in a model that included all three event types (t(67)=1.18, p=.24). No significant effect of interaction was found. Similarly, no results were found for the alternative measures of correlation of daily average tic rate in tic-triggering events (CONDITION: t(67)=0.77, p=.44, ORDER: t(67)=1.88, p=.06 for all events, and CONDITION: t(67)=1.07, p=.29, ORDER: t(67)=1.04, p=.3 for BATTLE and CHEST) and change from the first to last days (CONDITION: t(67)=1.07, p=.28, ORDER: t(67)=1.23, p=.22 for all events, and CONDITION: t(67)=0.54, p=.59, ORDER: t(67)=0.94, p=.35 for BATTLE and CHEST). No interaction was found in any of these cases.

### Pre-to-post-XTics change in tic severity, urge, and comorbidity measures

As indicated in Table S1, correlations among the three tic severity measures – YGTSS, PTQ, and Rush – were only partially observed (we found no significant correlation between PTQ and Rush at any of the assessed time points). However, in line with H8, a common reduction in tic severity was observed following XTics training across all three assessment measures: YGTSS tic severity scores, PTQ, and Rush (see Figure 3a and Table 3 for more details). Notably, PTQ scores remained significantly lower than their baseline levels (mean individual change of 42.99±31.69%, Z=-4.38, p=6×10^-6^) three months after the completion of the XTics training. XTics demonstrated a decrease in YGTSS tic severity scores also when compared to both control Group-EIT (Z=-5.71, p=6×10^-9^) and Group-CBIT (Z=-4.68, p=2×10^-6^) groups after 8 sessions as reported in ^32^. Similarly, the reduction in PTQ scores following XTics was significantly greater that in Group-EIT (Z=-3.9, p=4×10^-5^) and Group-CBIT (Z=-1.7, p=.04). These effects remained significant after applying FDR correction for multiple comparisons within H11 and H12.

**Figure 3:**
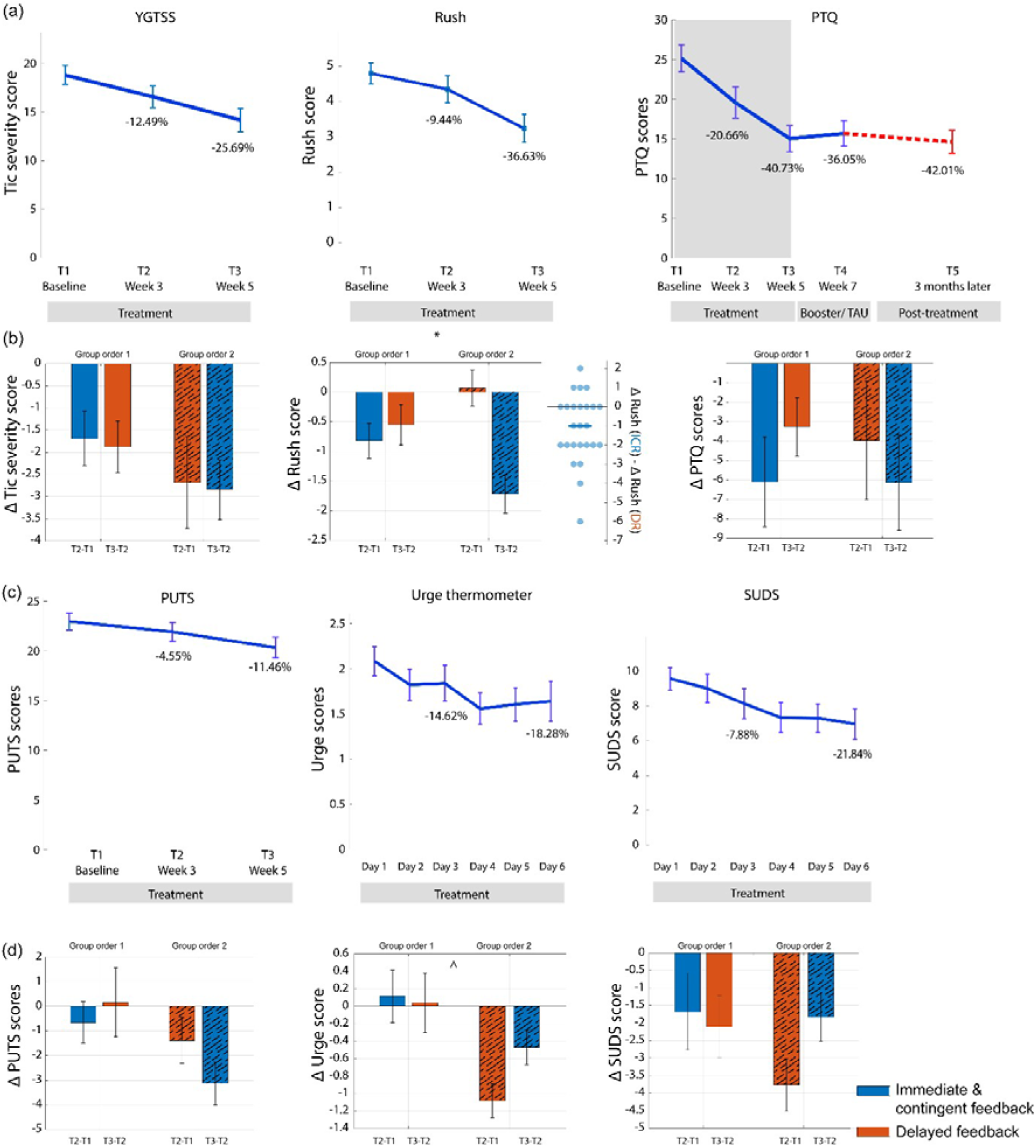
Change in tic severity during and after XTics training. (a) Time courses of the tic severity measures. The numbers represent the average group score differences from the baseline (unlike Table 3, which displays the average individual changes from baseline). (b) Differences in tic severity measures before and after each of the two alternative XTics phases. In the case of Rush, swarm plot of comparing score differences between the conditions is introduced. The blue horizontal line represents the median value. * CONDITION effect at Q_FDR_<.05. (c) and (d) are similar to (a) and (b), but with measures of tic-related urge and inconvenience.

**Table 3:**
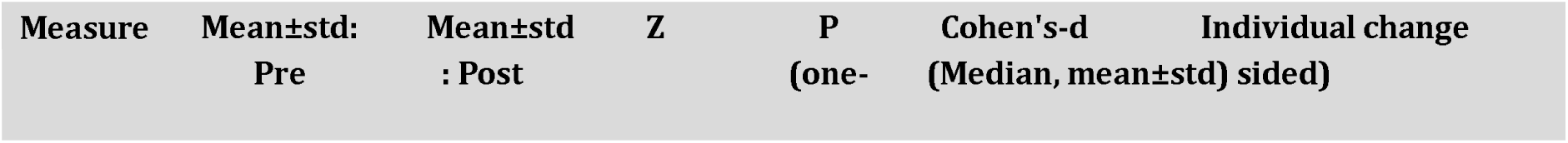
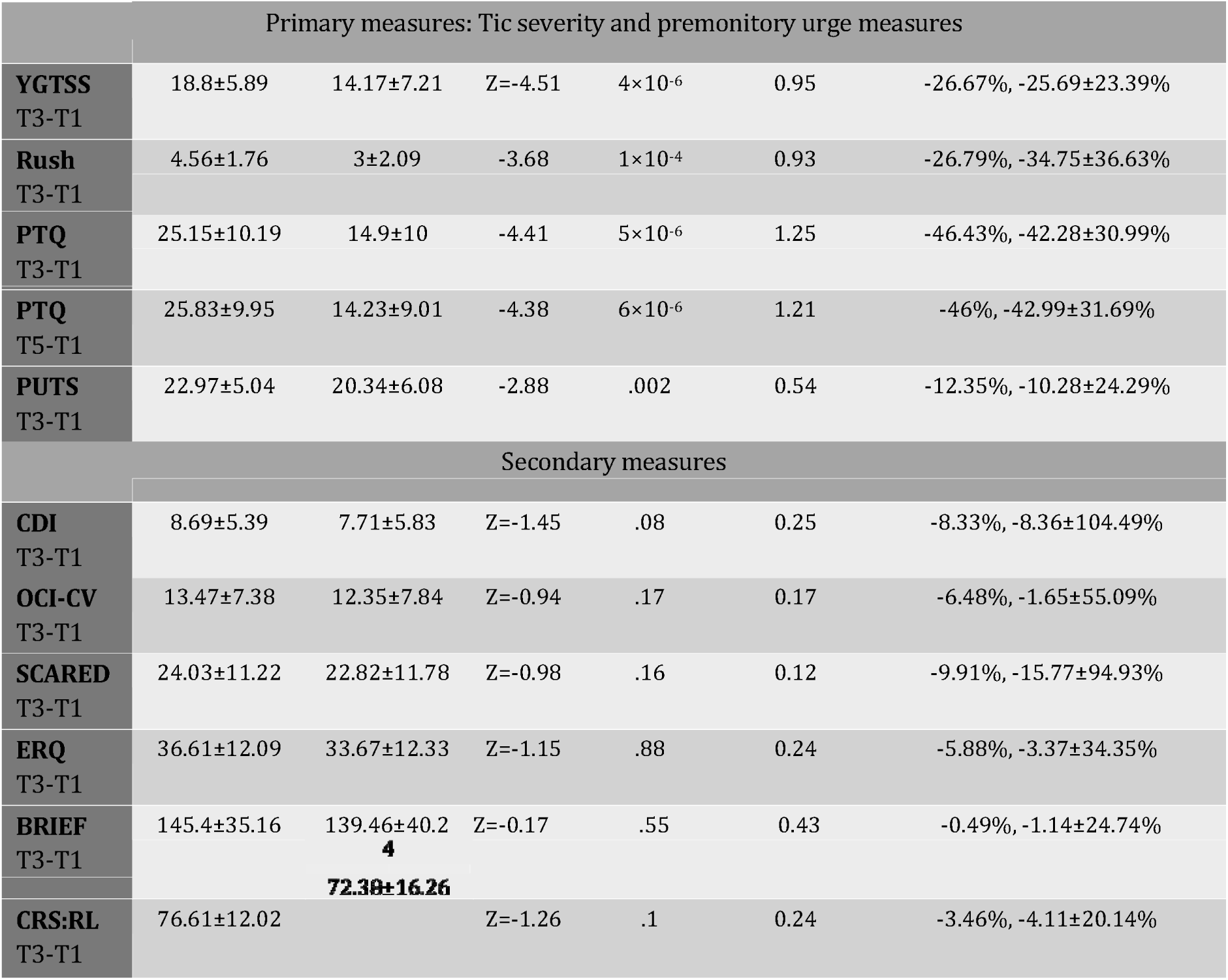
Pre-Post XTics changes in tic severity, premonitory urge and comorbid symptoms. T1 – Baseline, T3 – Week 5 (completion of the XTics clinical protocol), T5 – three months post-completion of the XTics clinical protocol. * The reason for the difference between these values is that the averages are calculated only for participants whose parents completed the PTQ questionnaire at both time points, and these differ between T3 and T5. Abbreviations: YGTSS - Yale Global Tic Severity Scale, PTQ - The Parent Tic Questionnaire, PUTS - Premonitory Urge for Tic Scale.

Moreover, in accordance with H9, there was a decrease in self-reported premonitory urge intensity and the associated distress related to premonitory urges and tics, as measured by SUDS, throughout the XTics training. Alongside a decline in PUTS scores (Table 3, Figure 3c), we found a daily reduction in these measures that remained consistent across two alternative sampling methodologies. We identified daily decrease in urge-thermometer scores (Z=-1.95, p=.026) and SUDS (Z=-4.32, p=8×10^-6^), as well as a significant reduction in scores between the maximal values on Day 1/2 and Day 6 for urge thermometer (t(31)=-3.06, p=.002) and SUDS (t(31)=-6.37, p<1×10^-5^). All results survived FDR correction within H9 and H10 at Q_FDR_<.05. The reduction in PUTS scores was significantly greater when compared to the corresponding CBIT group (Z=-3.56, p=.0002), but not when compared to a control psychoeducation group (Z=-0.33, p=.37).

On the other hand, no significant pre-post XTics change was observed in any of the measures of comorbid symptoms of depression, obsessive-compulsive disorder, anxiety, emotion regulation, and executive functions (Table 3). In an exploratory analysis, no effect was found either in the ERQ subscales of reappraisal (Z=-1.45, p=.93) and suppression (Z=-1.3, p=.9). The improvement in comorbidity symptoms was insignificant also when compared with both Group-EIT (CDI: Z=.46, p=.68; SACRED: Z=0.89, p=.81; ERQ: Z=-0.71, p=.76) and Group-CBIT (CDI: Z=.07, p=.53; SACRED: Z=2.08, p=.98; ERQ: Z=-0.72, p=.76) scores.

### The effect of the gaming feedback mode on tic severity and premonitory urge measures

As depicted in Figure 3b, our findings diverge from H15, as we did not observe a significant main effect of CONDITION on the tic severity measures represented by YGTSS (t(67)=0, p=.98) and PTQ (t(57)=1.08, p=.42). Neither ORDER nor interaction was found for YGTSS (t(67)=1.2, p=.24; t(66)=0.24, p=.82, respectively) and PTQ (t(57)=0.15, p=.8; t(56)=-0.14, p=.9). Nevertheless, in accordance with H15, we did identify a notable main effect of CONDITION on Rush scores (t(47)=3.47, p=.002, Q_FDR_=.03). This outcome suggests that tic severity exhibited a greater reduction following ICR as compared to DR. ORDER and interaction were not significant for Rush (t(47)=0.43, p=.6; t(46)=2.46, p=.045, Q_FDR_>.05).

No significant CONDITION effect was observed for changes in self-reported premonitory urge intensity (t(65)=1.32, p=.3), urge-related distress measured via thermometer scale (Pearson: t(61)=0.28, p=0.7; difference: t(61)=1.63, p=0.11), or SUDS (Pearson: t(61)=-1.91, p=.08; difference: t(61)=-1.49, p=.16).

A significant main effect of ORDER emerged in the analysis of post-pre differences in urgerelated distress using the thermometer scale (t(61)=3.37, p=.003, Q_FDR_=.03). Specifically, the group receiving ICR before DR exhibited a greater reduction in scores on this scale. However, an alternative Pearson measurement of the same effect did not survive correction for multiple comparisons (t(61)=2.26, p=.03, Q_FDR_=.16). No significant ORDER effect was observed for the urge measures PUTS (t(65)=2.02, p=.01, Q_FDR_=.12) and SUDS (Pearson: t(61)=-0.26, p=.84; difference: t(61)=1.06, p=.32), nor for the tic severity measures YGTSS (t(67)=1.23, p=.24), PTQ (t(57)=0.16, p=.8), and Rush (t(47)=.43, p=.6).

Significant CONDITION×ORDER interactions were not observed for any of the measures: YGTSS (t(66)=0.24, p=.82), PTQ (t(56)=-0.15, p=.91), Rush (t(46)=2.46, p=.05, Q_FDR_=.21), PUTS (t(64)=0.48, p=.71), urge thermometer (Pearson: t(60)=1.02, p=.31; difference: t(60)=1.13, p=.28), and SUDS urge thermometer (Pearson: t(61)=0.79, p=.44; difference: t(61)=0.94, p=.37).

### Alterations in tic-to-tic intervals and changes in tic severity following booster training

Consistent with H17, a notable increase in TTTI occurred during the 2-day booster training (=0.3±0.27, z=3.63, p=.0002). There were no significant differences in PTQ scores between the ratings before (T3) and after (T4) the booster/home-training phase (Z=-0.02, p=.98), nor when comparing T4 with scores obtained three months later (T5; Z=-0.96, p=.33). Contrary to H18, the difference in PTQ scores between T4 and T5 was not significantly greater in the group that underwent the booster training (Z=0.68, p=.5). Additionally, no differences were found between groups when comparing T4 and T3 (Z=-0.21, p=.84), and T5 and T3 (Z=0, p=1).

### Correlation of tic severity change with demographic, personality, and performance measures

Table 5 presents the results of tests evaluating the relationships between post-pre XTics and ICR phase changes in tic severity measures, and the factors gender, age, ADHD, emotion regulation skills, user engagement, executive functions, and changes in TTTI during training. While none of the effects remained significant after adjusting for multiple comparisons, it is noteworthy that there was evidence of a greater reduction in Rush scores for males compared to females post-pre XTics (Z=2.32, P=.02) and post-pre ICR (Z=2.66, P=.008). Additionally, a negative correlation was observed between the change in Rush scores post-pre XTics and TTTI AUC (R=-0.44, p=.02). Interestingly, TTTI AUC also showed a trend towards a negative correlation with the change in PTQ scores three months post-training relative to baseline (R=0.36, p=.05). However, these findings should be interpreted with caution, as they do not remain significant after correction for multiple comparisons.

**Table 5:**
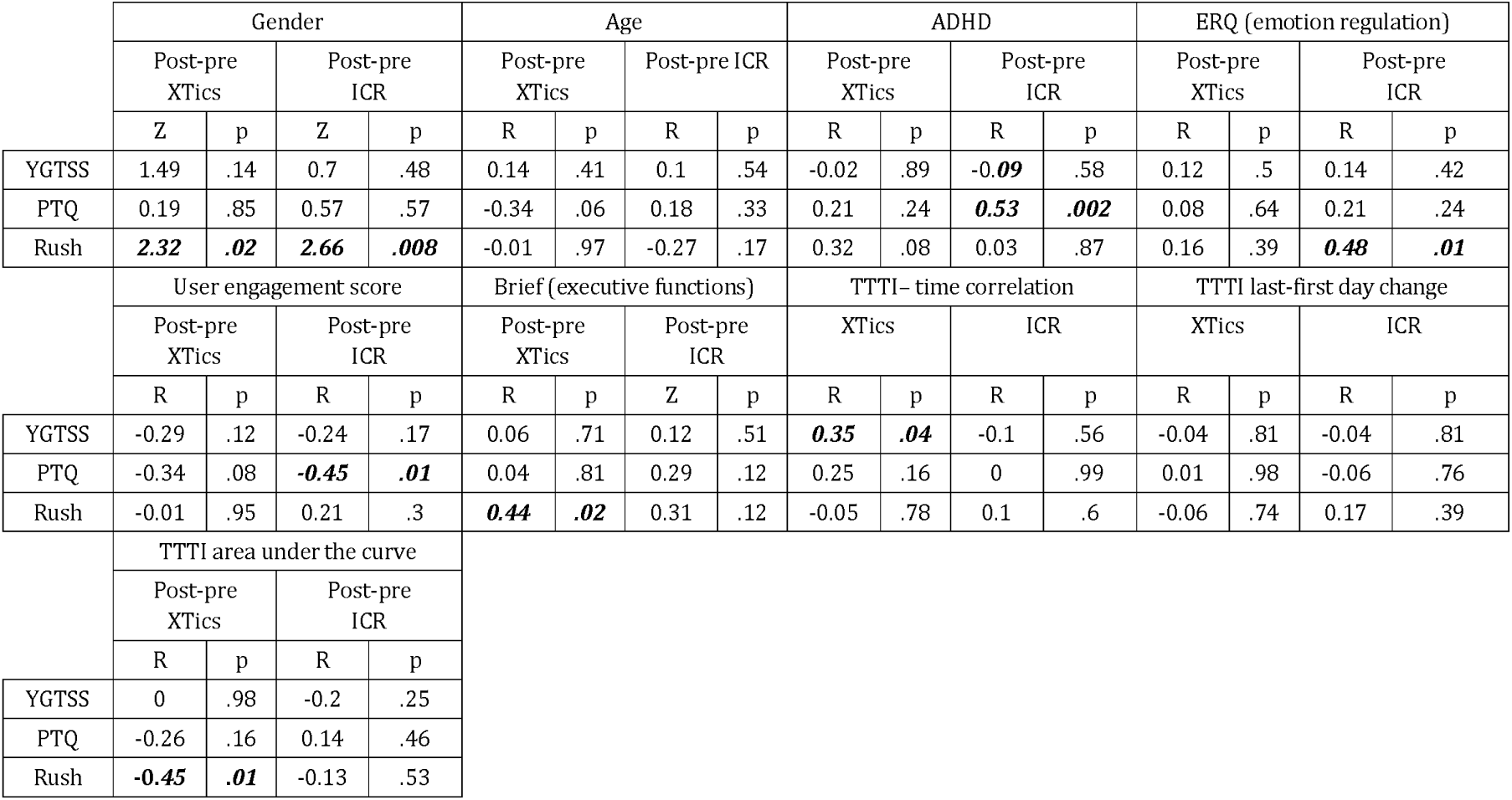
Correlations between changes in tic severity measures and demographic, personality, and game performance measures. Correlations with p<.05 are emphasized.

|  | Gender |  |  |  | Age |  |  |  | ADHD |  |  |  | ERQ (emotion regulation) |  |  |  |
| --- | --- | --- | --- | --- | --- | --- | --- | --- | --- | --- | --- | --- | --- | --- | --- | --- |
|  | Post-pre XTics |  | Post-pre ICR |  | Post-pre XTics |  | Post-pre ICR |  | Post-pre XTics |  | Post-pre ICR |  | Post-pre XTics |  | Post-pre ICR |  |
|  | Z | p | Z | p | R | p | R | p | R | p | R | p | R | p | R | p |
| YGTSS | 1.49 | .14 | 0.7 | .48 | 0.14 | .41 | 0.1 | .54 | -0.02 | .89 | -0.09 | .58 | 0.12 | .5 | 0.14 | .42 |
| PTQ | 0.19 | .85 | 0.57 | .57 | -0.34 | .06 | 0.18 | .33 | 0.21 | .24 | 0.53 | .002 | 0.08 | .64 | 0.21 | .24 |
| Rush | 2.32 | .02 | 2.66 | .008 | -0.01 | .97 | -0.27 | .17 | 0.32 | .08 | 0.03 | .87 | 0.16 | .39 | 0.48 | .01 |
|  | User engagement score |  |  |  | Brief (executive functions) |  |  |  | TTTI-time correlation |  |  |  | TTTI last-first day change |  |  |  |
|  | Post-pre XTics |  | Post-pre ICR |  | Post-pre XTics |  | Post-pre ICR |  | XTics |  | ICR |  | XTics |  | ICR |  |
|  | R | p | R | p | R | p | Z | p | R | p | R | p | R | p | R | p |
| YGTSS | -0.29 | .12 | -0.24 | .17 | 0.06 | .71 | 0.12 | .51 | 0.35 | .04 | -0.1 | .56 | -0.04 | .81 | -0.04 | .81 |
| PTQ | -0.34 | .08 | -0.45 | .01 | 0.04 | .81 | 0.29 | .12 | 0.25 | .16 | 0 | .99 | 0.01 | .98 | -0.06 | .76 |
| Rush | -0.01 | .95 | 0.21 | .3 | 0.44 | .02 | 0.31 | .12 | -0.05 | .78 | 0.1 | .6 | -0.06 | .74 | 0.17 | .39 |
|  | TTTI area under the curve |  |  |  |  |  |  |  |  |  |  |  |  |  |  |  |
|  | Post-pre XTics |  | Post-pre ICR |  |  |  |  |  |  |  |  |  |  |  |  |  |
|  | R | p | R | p |  |  |  |  |  |  |  |  |  |  |  |  |
| YGTSS | 0 | .98 | -0.2 | .25 |  |  |  |  |  |  |  |  |  |  |  |  |
| PTQ | -0.26 | .16 | 0.14 | .46 |  |  |  |  |  |  |  |  |  |  |  |  |
| Rush | -0.45 | .01 | -0.13 | .53 |  |  |  |  |  |  |  |  |  |  |  |  |

## Discussion

Our study supports the notion that ERP treatment for tic disorders can be significantly enhanced by a varied gamified (presumably dopaminergic) context, paired with immediate reinforcement. First, our gamified training protocol achieved near-complete compliance and reported high user engagement. This outcome is notable, especially considering previous reports where half of the children undergoing behavioral tic therapy dropped out after completing only half of the recommended number of sessions^18^.

Second, consistent with our expectations, the interface version featuring only stimulating events with delayed feedback on tic suppression effectively trained participants in the key ERP task for TDs: extending tic-free intervals. Additionally, the XTics version that supplemented the stimulating events with immediate and contingent feedback led to even greater improvement in this skill. Notably, this effect was more pronounced in later training sessions, particularly when the ICR version was introduced only in the second training week. These findings indicate that extending the TTTI is a skill learned gradually, where immediate feedback becomes especially beneficial after achieving a basic level of mastery.

The findings indicate that our gamified intervention was effective not only in training participants to extend the TTTI but also in reducing real-life tic severity, along with the intensity and inconvenience of related urges. After six training sessions, all three measures of tic severity significantly decreased. This reduction was notably greater than the effects observed in comparable cohorts undergoing psychoeducation and group CBIT. Notably, 54.9% of our participants showed more than a 25% reduction in total tic severity score, a threshold identified as indicating clinically significant change^49^.

Figure S2 provides an anecdotal comparison of the post-pre YGTSS reduction observed in our study with effects reported in other studies using cognitive-behavioral interventions for TD. The average YGTSS reduction in XTics was less than one composite standard deviation below the mean in only one of seven cases^11^, where a longer (10 weeks) and more intensive training protocol (including 15 minutes of daily practice) was employed. In two other instances, and in all comparisons with control protocols (waitlist, psychoeducation), the mean YGTSS reduction exceeded one composite standard deviation above the corresponding measure. This indicates that XTics achieves an impact comparable to other cognitive-behavioral interventions, despite being measured after only five weeks rather than the typical ten. Notably, XTics was designed to replace the ‘homework component’ of ERP. The relative efficacy of XTics, despite the absence of this crucial component, highlights its potential as a beneficial addition to existing treatments for TDs.

As anticipated, we observed that the ICR condition, combining tic stimulation with immediate feedback, resulted in a greater reduction in Rush scores compared to the DR condition. This suggests that the contingency and immediacy of feedback on tic suppression significantly enhance clinical outcomes. However, there were no significant differences between ICR and DR in our other tic severity measures - YGTSS and PTQ. It is important to note that YGTSS and PTQ assess persistent and significant changes in tic manifestation characteristics (for example, a reduction of only 1 YGTSS point occurs when decreasing the number of tics from 2-5 to 1). Additionally, while YGTSS and PTQ are often used in studies to evaluate changes in tic severity accumulated over several weeks or months, we assessed changes over the shortest time-frame these measures cover: one week. Therefore, longer intervention periods may be necessary to detect the impact of ICR on YGTSS and PTQ, as these scales have lower temporal sensitivity compared to Rush.

Our study found only limited evidence, not robust enough to withstand correction for multiple comparisons, that a greater reduction in tic severity correlates with higher success in elongating TTTI (as indicated by the TTTI AUC). To confirm this finding, it should be replicated in future studies. The relationship between TTTI elongation and reduced tic severity might be complicated by various factors that obscure their correlation. For instance, several participants achieved maximal baseline TTTI while playing ZenithX, indicating that their further clinical improvement could not correspond with an increase in TTTI. Apart from this potential ceiling effect, the correlation between TTTI elongation and tic severity measures might be skewed by factors affecting the translation of learned tic suppression skills to real-life situations. These mediating factors, not the focus of our current study, should be the subject of future research.

It should be noted that we observed no effect of the treatment on comorbid symptoms. This could be attributed to the short-term intervention’s lack of focus on these comorbidities, which typically necessitate comprehensive behavioral therapy.

### Caveats and Limitations

Beyond the limitations of the crossover design in assessing long-term clinical effects and the game’s ineffectiveness in providing significant feedback when the TTTI extends to several minutes, we identified other caveats. Firstly, although ZenithX was initially engaging, sustained user interest might wane due to its basic game design. Future versions could address this by diversifying the challenges and narrative, while preserving crucial elements like reward anticipation and immediate feedback. Additionally, while the current study examined XTics in a population of children aged seven years and above with moderate to high tic severity, the efficiency of the protocol should be tested on other populations. Finally, XTics’ dependence on manual tic detection limits its scalability and poses challenges in terms of reliability. Integrating validated computer vision-based tic detection^50^ could enhance XTics’ practicality and reliability.

## Conclusion

Our study supports the concept that tic triggering and incremental enhancement through immediate, contingent gamified feedback on tic frequency effectively improve tic suppression skills. While exposure and delayed feedback are beneficial, the addition of rapid feedback significantly boosts this effect. The incorporation of such a gamified platform into behavioral interventions for TD, especially for children, may enhance both treatment compliance and efficacy.

## Data Availability

All data produced in the present study are available upon reasonable request to the authors under the hospital's constraints

## Acknowledgements

We thank Professor Talma Hendler, the Founding Director of the Sagol Brain Institute at Tel-Aviv Sourasky Medical Center, for her valuable support, Maya Zuckerman and our partners at Actiview: Eran Ravid, Matanel Libi, Shani Libi, Gil Asher, and Lanir Ben Shoshan for collaborating in the development of ZenithX, and Yael Abarbanel who was part of the clinical staff.

