## Supplementary Material for "Gamified closed-loop non-pharmacological intervention enhances tic suppression in children"

**ZenithX algorithm**

In the immediate and contingent reward (ICR)mode of ZenithX, we calculate a baseline average TTTI (ATTTI), and an average gap (AG) between successive TTTIs that are equal or higher than the ATTTI. During the game,

**Research team training**

Clinician training

Four clinicians, including two advanced BA psychology students, an MA psychology student, and a social work practitioner, underwent training in behavioral therapy (Exposure and Response Prevention - ERP) for tic disorders under the guidance of senior cognitive-behavioral therapist SZB. The clinicians gained a comprehensive understanding of the negative reinforcement hypothesis of tic maintenance, the rationale and fundamentals of ERP, and various relaxation strategies. Subsequently, the trainees received instruction in the administration of clinical assessment tools.

Co-therapist training

Similarly, five co-therapists were familiarized with the negative reinforcement hypothesis of tic maintenance, the rationale and fundamentals of ERP, and various relaxation strategies. Subsequently, they underwent a training program for real-time tic coding led by two experienced research associates. Utilizing a tic coding method developed by our research team for a preliminary study[1], the co-therapists coded videotaped data from subjects who participated in that study to acquire proficiency in identifying and categorizing tics as either vocal or motor, simple or complex (based on the YGTSS responses of each subject). The trainee co-therapists coded 10-minute segments of audiovisual data by employing assigned keys on the computer keyboard. The coding output was then compared to the output obtained by an experienced research associate.

Upon completion of the treatment program, we implemented an additional coding procedure on eight selected game rounds, which were videotaped and recorded during the exercise meetings of four subjects. Each co-therapist coded a game round recording conducted by a different co-therapist and disagreements were discussed.

**Virtual introduction to ERP**

The virtual group 90 minute sessions were performed via Zoom separately to participants of younger (7-10 years) and older (11-15 years) age by a senior cognitive-behavioral therapist (SZB) with expertise in this treatment. These sessions focused on psychoeducation on tics and ERP principles and also included the acquisition of relaxation techniques such as deep diaphragmic breathing and progressive muscle relaxation.

**Changes from the pre-registered analysis plan**

Inadvertently, the pre-registration outlined a repeated measures ANOVA to assess inter-condition differences. However, this approach proved infeasible due to the presence of missing values. Consequently, we opted for a mixed linear model, capable of accommodating such instances, to analyze both between-subject effects (ORDER) and within-subject effects (CONDITION).

In the pre-registration, we declared that Hypothesis H8 will be tested using a paired test, but with the resulting t-statistic compared to a null distribution of 100,000 values generated by shuffling the time labels. This test resulted in a highly significant effect: $t(30)=-4.99$, $p<1\times10^{-5}$ (i.e., none of the tests of the reshuffled data resulted in a more extreme t value). However, since we now see no reason to test this measure differently from the YGTSS and PTQ scores, we similarly report on the outcomes of a Wilcoxon test for the Rush scores.

Similarly, in Hypothesis H15, we replaced Mann-Whitney test with a permutation test and tested the impact of CONDITION and ORDER on SUDS and premonitory urge intensity examining the difference between the final scores and the maximal scoring in day 1 and day 2 of each phase as in H9.

The Conner's Parent Rating Scale-Revised Long Form (CRS: RL) and Obsessive Compulsive Inventory–Child Version (OCI-CV) data were not available for the control groups Group-CBIT and Group-EIT[4] so they were removed from H13, H14.

| **T0** | | | | **T1** | | | **T2** | | |
|---|---|---|---|---|---|---|---|---|---|
| | **YGTSS** | **PTQ** | **Rush** | **YGTSS** | **PTQ** | **Rush** | **YGTSS** | **PTQ** | **Rush** |
| **YGTSS** | 1 | 0.39* | 0.44* | 1 | 0.64*** | 0.5* | 1 | 0.71*** | 0.37^ |
| **PTQ** | 0.39* | 1 | 0.17 | 0.64*** | 1 | 0.28 | 0.71*** | 1 | 0.23 |
| **Rush** | 0.44* | 0.17 | 1 | 0.5* | 0.28 | 1 | 0.37^ | 0.23 | 1 |

**Table 4**: Spearman's Correlation between tic severity measures at three time points. ^$Q_{FDR}$ =.06, * $Q_{FDR}$ <=.05, ** $Q_{FDR}$<=.005, *** $Q_{FDR}$<=.0005

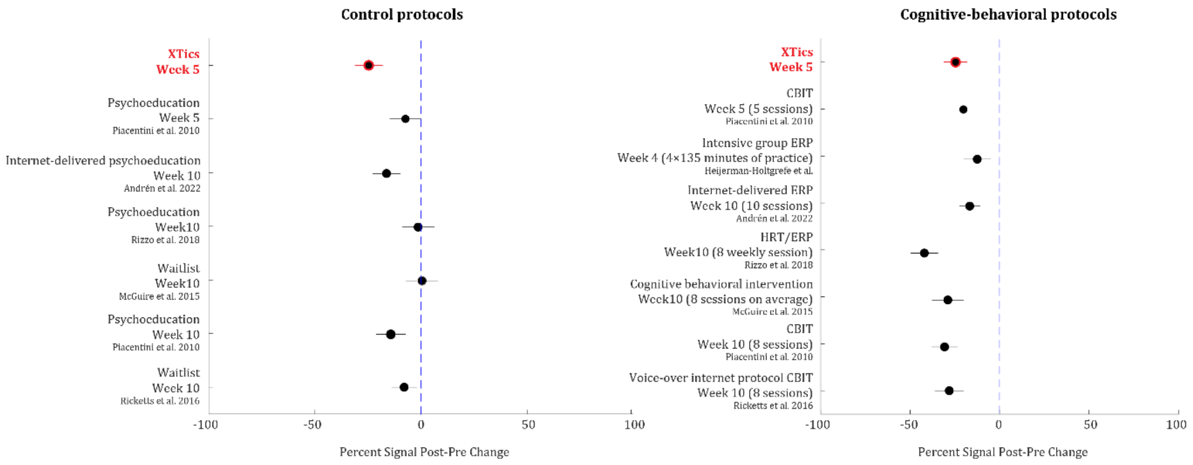


Figure S2: Forest plot comparing pre- and post-intervention changes in YGTSS Scores in XTics against findings from other studies. This comparison includes studies identified in a recent meta-analysis examining the efficacy of cognitive behavioral therapies for tic disorders[5]. Selection criteria included studies specifically reporting YGTSS outcomes and involving age groups comparable to our study. Additionally, we incorporated findings from two recent ERP studies by [6,7], which were not included in the original meta-analysis.

The plot visualizes the treatment effect as a standardized percent signal change for each study, with the variability represented using a composite standard deviation calculated from both reported baseline and post-treatment measurements. The composite standard deviation is computed as $\sqrt{\frac{Basline\ Std^2 + Post-treatment\ Std^2}{2}}$. Each point represents the mean percent change for a study, with horizontal lines indicating the variability range based on the composite standard deviation, providing a comparative view of treatment effectiveness across studies. Disclaimer: The analysis does not account for intra-individual variability due to the unavailability of individual within-subject differences in the reports. As such, while providing valuable insights, this approach might be suboptimal compared to methods that directly analyze within-subject changes. Included studies: [7–12]


1. Raz, G. *et al.* Impact of movie and video game elements on tic manifestation in children. *Eur. J. Neurol.* **31**, e16120 (2024).
2. Silverman, W. K. & Albano, A. M. *Anxiety disorders interview schedule for DSM-IV: Child version*. (Oxford University Press, 1996).
3. Hirschtritt, M. E. *et al.* Lifetime Prevalence, Age of Risk, and Genetic Relationships of Comorbid Psychiatric Disorders in Tourette Syndrome. *JAMA Psychiatry* **72**, 325–333 (2015).
4. Zimmerman-Brenner, S. *et al.* Group behavioral interventions for tics and comorbid symptoms in children with chronic tic disorders. *Eur. Child Adolesc. Psychiatry* **31**, 637–648 (2022).
5. Shou, S. *et al.* The Efficacy of Cognitive Behavioral Therapy for Tic Disorder: A Meta-Analysis and a Literature Review. *Front. Psychol.* **13**, 851250 (2022).
6. Andrén, P. *et al.* Therapist- and parent-guided Internet-delivered behaviour therapy

for paediatric Tourette's Disorder: a pilot randomised controlled trial with long-term follow-up. doi:10.31234/OSF.IO/DP3QZ
